# Impact Of Maternal Education on Perinatal Outcomes in Delta State, Nigeria

**DOI:** 10.64898/2026.04.30.26352195

**Authors:** Mordecai Oweibia, Gift Cornelius Timighe, Ebiakpor Bainkpo Agbedi

## Abstract

**Background:** Perinatal mortality remains a major public health concern in Nigeria despite global progress in maternal and child health. Maternal education has been identified as a key determinant influencing perinatal outcomes through its effects on health literacy, service utilization, and decision-making. However, limited evidence exists on how maternal education directly impacts perinatal outcomes within the context of Delta State, Nigeria. This study therefore investigated the relationship between maternal education and perinatal outcomes, focusing on perinatal mortality, access to healthcare, and educational interventions that enhance maternal health.

**Methods:** A **quantitative cross-sectional study design** was employed. Data were collected from **400 mothers** who delivered in selected public and private health facilities across six Local Government Areas in Delta State, alongside secondary data on perinatal outcomes obtained from hospital records. A **structured questionnaire** and **record extraction form** were used to gather information on maternal education, healthcare access, and perinatal indicators. Data were analyzed using **SPSS Version 26**, applying descriptive statistics, Pearson’s correlation, and regression analysis to determine associations between maternal education and perinatal outcomes.

**Results:** Findings revealed a strong inverse relationship between maternal education and perinatal mortality (**r = −0.431, p < 0.01**), indicating that mothers with higher education levels experienced fewer stillbirths and neonatal deaths. Similarly, maternal education was significantly associated with reduced low birth weight incidence (**r = −0.362, p < 0.01**) and improved neonatal survival (**r = 0.415, p < 0.01**). Regression results showed that maternal education accounted for **23.9% of the variance** in perinatal outcomes (**R**^**2**^ **= 0.239, p < 0.001**). Women with tertiary education were more likely to attend antenatal care (94%), deliver in health facilities (91%), and receive postnatal care (89%) compared to those without formal education.

**Conclusion:** The study concludes that maternal education plays a decisive role in improving perinatal outcomes in Delta State by promoting healthcare utilization, enhancing health literacy, and reducing preventable perinatal deaths. Strengthening women’s education through formal schooling and community-based literacy programs is vital for achieving equitable maternal and neonatal health outcomes. The study recommends multisectorial collaboration between education and health authorities to integrate maternal health education into national curricula and community outreach initiatives as part of efforts to attain **Sustainable Development Goals 3 and 4**.

## INTRODUCTION

### 1.0 Introduction

Maternal education has long been recognized as a powerful determinant of health and social development outcomes, shaping the well-being of both mothers and their infants. In public health literature, the educational level of mothers is often cited as one of the most influential predictors of perinatal health encompassing the period from pregnancy to the first week of life. The significance of maternal education stems from its profound influence on health-seeking behavior, decision-making capacity, nutritional choices, and adherence to medical advice. These elements collectively contribute to maternal and child survival, especially in low- and middle-income countries where health inequities remain pronounced (Kozuki et al., 2019; AbouZahr et al., 2021; Oweibia et al., 2024; Oweibia et al., 2026).

Globally, the World Health Organization (WHO) estimates that approximately 2 million perinatal deaths occur annually, with a large proportion concentrated in sub-Saharan Africa (WHO, 2023). Within this context, maternal education plays a pivotal role in reducing preventable perinatal deaths through enhanced awareness, timely utilization of antenatal and delivery services, and improved home-care practices (Bhowmik et al., 2022). Studies have shown that mothers with higher education levels are more likely to access skilled birth attendants, a pattern consistently observed in Nigeria’s maternal and child health trends (Mordecai et al., 2025; Oweibia et al., 2026). Women with higher education levels are more likely to deliver in health facilities, and adopt safer postnatal practices, ultimately improving perinatal survival rates (Ali et al., 2020; Adebowale & Adedini, 2021).

In Nigeria, perinatal mortality remains a critical public health concern, reflecting disparities in access to quality healthcare and maternal literacy. The National Demographic and Health Survey (NDHS, 2021) reports that the perinatal mortality rate is approximately 39 per 1,000 live births, with significant regional variations. Delta State, despite being one of Nigeria’s more socioeconomically active regions, continues to face challenges in maternal and perinatal health outcomes. Factors such as maternal illiteracy, low socioeconomic status, and limited awareness of available healthcare services contribute to these outcomes (Ntoimo & Okonofua, 2020).

Educational attainment enhances a mother’s ability to comprehend health information, communicate effectively with healthcare providers, and recognize complications early enough to seek intervention (Kassaw et al., 2023). Consequently, education empowers women to make informed decisions regarding antenatal attendance, delivery location, and neonatal care all of which are integral to perinatal health outcomes. Conversely, lack of education often correlates with high-risk behaviors, delayed care-seeking, and limited knowledge of preventive measures.

This study is therefore motivated by the persistent perinatal health disparities observed in Delta State and the broader Nigerian context. It seeks to analyze how varying levels of maternal education influence perinatal outcomes such as neonatal mortality, stillbirths, and low birth weight. In addition, the study explores the role of educational interventions in promoting maternal health and identifies the extent to which maternal education facilitates access to healthcare services.

### 1.1 Background of the Study

Maternal education remains one of the most powerful predictors of perinatal health outcomes globally. The perinatal period extending from the 22nd week of gestation to seven days after birth is a highly sensitive stage where both maternal and neonatal health risks are interlinked. Education equips mothers with knowledge, skills, and attitudes necessary for making informed health decisions that can mitigate perinatal risks (Kozuki et al., 2019; Kassaw et al., 2023). From a public health perspective, educational attainment enables women to understand health information, access healthcare services promptly, and apply preventive and curative interventions effectively. Consequently, improving maternal education is widely acknowledged as a key driver with empirical support from national analyses demonstrating improvements in maternal and neonatal indicators over time (Mordecai et al., 2025). Overtime women education has brought about sustainable reductions in perinatal mortality and morbidity rates (World Health Organization [WHO], 2023).

Globally, perinatal outcomes have improved markedly over the last two decades due to increasing investments in maternal and child health, yet wide disparities persist between developed and developing nations. According to WHO (2023), sub-Saharan Africa accounts for nearly 43% of global perinatal deaths, largely due to preventable causes such as infections, birth asphyxia, and preterm complications. Studies indicate that mothers with secondary or tertiary education levels have significantly better perinatal outcomes compared to those with no formal education, owing to enhanced awareness and healthcare utilization (Bhowmik et al., 2022; AbouZahr et al., 2021). Education not only improves knowledge of antenatal and delivery care but also increases household income, enabling better nutrition and access to skilled birth attendants.

In Nigeria, the link between maternal education and perinatal outcomes is particularly pronounced. Despite policy efforts such as the National Strategic Health Development Plan (NSHDP II, 2018–2022), the country still records one of the world’s highest perinatal mortality rates approximately 39 deaths per 1,000 live births (National Population Commission [NPC] & ICF, 2021). Socioeconomic inequalities and gender disparities exacerbate these outcomes. Women with little or no formal education are less likely to attend antenatal clinics, deliver in hospitals, or recognize danger signs during pregnancy (Adebowale & Adedini, 2021). Conversely, educated mothers are better positioned to understand medical instructions, plan for safe deliveries, and access family planning services. A study by Ali et al. (2020) found that in Nigeria, each additional year of maternal education reduces the probability of perinatal death by nearly 9%, highlighting education’s measurable impact on neonatal survival.

Delta State, one of Nigeria’s oil-producing regions, represents a complex socioeconomic context where educational disparities coexist with relatively advanced healthcare infrastructure. Despite having multiple health facilities, multiple public health interventions aimed at improving maternal and child health outcomes, the state continues to experience high rates of perinatal complications including low birth weight and stillbirths (Oweibia et al., 2023; Gabriel et al., 2023; Ntoimo & Okonofua, 2020). Contributing factors include limited maternal literacy, poverty, and cultural practices that discourage women from seeking institutional care. Many rural and semi-urban communities in Delta State rely on traditional birth attendants, often due to misinformation or lack of awareness about the benefits of skilled healthcare. Educational interventions aimed at empowering women, such as community literacy programs and antenatal education initiatives have shown promise in improving maternal and neonatal outcomes in similar settings (Usman et al., 2022).

From a theoretical standpoint, the relationship between maternal education and perinatal outcomes can be explained through the *Social Determinants of Health (SDH)* framework, which posits that health inequities stem largely from social, economic, and educational inequalities. Education enhances cognitive skills and fosters autonomy, leading to improved health behaviors and increased utilization of medical services (Marmot & Allen, 2020). Historically, public health strategies dating back to the Alma-Ata Declaration of 1978 emphasized education as a cornerstone of primary healthcare, highlighting its role in empowering communities toward better health practices. Modern health promotion models, such as the Health Belief Model, further support the notion that perceived knowledge and self-efficacy both products of education drive health-seeking behaviors that reduce maternal and neonatal risks (Becker et al., 1974).

Despite progress, challenges persist in achieving equitable access to maternal education and healthcare in Nigeria. The intersection of poverty, cultural norms, and limited educational opportunities continues to undermine maternal and child health efforts. Moreover, while several national studies have demonstrated a general link between education and child survival, fewer have specifically examined *perinatal outcomes* a critical indicator of both maternal and neonatal well-being. Local-level analyses within Delta State remain limited, especially those integrating both quantitative data from health facilities and survey responses from mothers. Therefore, there is a pressing need for context-specific research that quantifies the impact of maternal education on perinatal health and identifies educational interventions that can effectively reduce perinatal mortality in the region.

### 1.2 Statement of the Problem

Despite global improvements in maternal and child health, perinatal mortality remains a pressing challenge in many developing countries, including Nigeria. The perinatal period is one of the most vulnerable stages of life, with outcomes heavily influenced by maternal knowledge, attitudes, and access to quality healthcare. While numerous global initiatives such as the Sustainable Development Goals (SDGs) and Nigeria’s National Health Policy have emphasized reducing maternal and neonatal deaths, progress has been uneven across regions. In Nigeria, particularly in Delta State, perinatal outcomes remain suboptimal, revealing persistent inequalities linked to maternal education and healthcare access.

Educational attainment among mothers plays a central role in shaping health behaviors, decision-making capacity, and utilization of maternal healthcare services. However, in Delta State, many women of reproductive age still possess low levels of formal education, especially in rural and semi-urban communities. This educational gap contributes to limited understanding of antenatal care schedules, delivery preparation, and early recognition of pregnancy complications. Consequently, low maternal literacy levels often result in delayed healthcare seeking, poor birth preparedness, and increased risk of perinatal mortality.

Furthermore, existing studies and public health interventions in Nigeria have primarily focused on general maternal and child health indicators, paying limited attention to the *specific pathways* through which maternal education influences perinatal outcomes. There remains a lack of empirical evidence at the state level particularly in Delta State linking educational attainment to measurable perinatal indicators such as stillbirths, neonatal deaths, and birth weight outcomes. This absence of localized, data-driven research hampers evidence-based policy formulation and the design of targeted educational interventions.

As highlighted in studies examining systemic barriers and health system performance in Nigeria, compounding these challenges are socioeconomic and infrastructural barriers that further diminish the effectiveness of maternal health programs (Oweibia et al., 2025; Agbedi & Oweibia, 2025). In several communities across Delta State, poverty, cultural beliefs, and limited health literacy hinder women’s ability to access modern healthcare facilities, even when such services are available. As a result, many deliveries still occur at home or with traditional birth attendants, thereby increasing the risk of perinatal complications and preventable deaths.

Given these challenges, there is a clear need to assess how maternal education affects perinatal outcomes within the specific sociocultural and economic context of Delta State. Understanding this relationship is crucial for designing interventions that not only improve women’s educational attainment but also enhance their utilization of maternal and child health services.

Therefore, this study seeks to bridge the existing knowledge gap by investigating the **impact of maternal education on perinatal outcomes in Delta State, Nigeria**, with particular emphasis on perinatal mortality rates, access to healthcare services, and the role of educational interventions in promoting maternal health. The findings will contribute to more targeted public health strategies aimed at reducing perinatal deaths and achieving equitable maternal health outcomes.

### 1.3 Objectives of the Study

The objectives of this study are structured to address the core research problem the persistent influence of maternal education on perinatal health outcomes in Delta State, Nigeria. This section outlines the general aim and specific objectives that guide the study’s direction and analysis.

#### General Objective

The main objective of this study is **to examine the impact of maternal education on perinatal outcomes in Delta State, Nigeria**. Specifically, the study aims to determine how varying levels of maternal education affect perinatal mortality, access to maternal healthcare services, and the effectiveness of educational interventions in improving maternal and neonatal health.

#### Specific Objectives

The study is designed to achieve the following specific objectives:

1. **To assess the level of maternal education in relation to perinatal death rates** among mothers who delivered in health facilities within Delta State.
2. **To identify specific educational interventions that improve maternal health** and contribute to better perinatal outcomes.
3. **To evaluate the correlation between maternal education and access to healthcare services** among women of reproductive age in Delta State.
4. **To analyze the role of socioeconomic and demographic factors** in moderating the relationship between maternal education and perinatal outcomes.
5. **To provide evidence-based recommendations** for policymakers and healthcare providers aimed at reducing perinatal mortality through educational empowerment programs.

### 1.4 Research Questions

Accordingly, the following research questions will guide this investigation:

1. **How does the level of maternal education relate to perinatal death rates** among mothers who delivered in health facilities in Delta State?
2. **What specific educational interventions contribute to improved maternal health** and better perinatal outcomes in Delta State?
3. **What is the correlation between maternal education and access to healthcare services** among women of reproductive age in Delta State?
4. **In what ways do socioeconomic and demographic factors moderate the relationship** between maternal education and perinatal outcomes?
5. **What policy measures or programs can be developed** to enhance maternal education and subsequently improve perinatal health indicators?

### 1.5 Scope of the Study

This study is geographically confined to **Delta State, Nigeria**, one of the South-South geopolitical regions of the country. Delta State is selected because it exhibits notable disparities in maternal education levels and perinatal health outcomes, despite having relatively developed healthcare infrastructure. The state includes both urban and rural populations, providing a balanced representation of differing socioeconomic and educational backgrounds. Data will be obtained from selected public and private healthcare facilities across various Local Government Areas (LGAs) to capture diverse maternal experiences.

The target population for this study comprises **mothers who delivered in healthcare facilities within Delta State** during the study period. The research focuses on women of reproductive age (15–49 years) who have experienced childbirth, as they are best positioned to provide relevant data on perinatal outcomes and healthcare utilization patterns. Healthcare professionals, including nurses and midwives, may also provide supplementary data on perinatal records to validate findings from survey responses.

The study covers a **five-year reference period (2018–2023)**, aligning with the most recent demographic and health data available in Nigeria. This timeframe allows for the assessment of trends in maternal education levels and perinatal outcomes while reflecting contemporary healthcare policies and educational interventions within the state.

Thematically, this study is limited to examining the relationship between **maternal education** and **perinatal outcomes** such as stillbirths, neonatal deaths, and low birth weight. It also investigates associated factors such as **access to healthcare services, health-seeking behaviors**, and **maternal literacy interventions**. Broader determinants of maternal and child health (e.g., environmental or genetic factors) are beyond the scope of this research and will not be analyzed in depth.

Methodologically, the study adopts a **quantitative cross-sectional design**, utilizing **survey data** collected from mothers and **secondary data** from healthcare facilities. Statistical analyses, including **descriptive statistics, correlation, and regression analysis**, will be employed to determine the strength and nature of relationships between variables. Data will be analyzed using recognized statistical software to ensure reliability and reproducibility of results. Ethical approval and informed consent procedures will also be strictly observed.

### 1.6 Significance of the Study

From an academic perspective, this study deepens scholarly understanding of the linkage between **education and maternal health outcomes** within the Nigerian context. While numerous global studies have examined the role of education in maternal and child health, relatively few have focused specifically on **perinatal outcomes** in subnational contexts such as Delta State. This research therefore fills a critical gap by providing localized empirical evidence that connects educational attainment to perinatal indicators, such as neonatal mortality and low birth weight. The findings will serve as a valuable reference for future researchers, scholars, and postgraduate students investigating maternal and child health determinants in similar settings.

The study also holds significant implications for **health policy and planning**. Evidence from this research can support the design and implementation of targeted educational interventions that enhance maternal literacy and healthcare awareness. Policymakers at both state and national levels can utilize the findings to refine existing maternal health programs including antenatal education initiatives, community outreach, and girl-child education policies. By highlighting the measurable impact of maternal education on perinatal outcomes, the study can inform policy frameworks aimed at reducing perinatal mortality rates and achieving national and international development goals such as the **Sustainable Development Goal (SDG) 3: Good Health and Well-being**.

For healthcare practitioners, particularly in Delta State, the study provides a basis for improving **maternal health education** within health facilities. Midwives, nurses, and community health workers can leverage the insights to design culturally appropriate communication strategies that resonate with women of varying literacy levels. Additionally, non-governmental organizations (NGOs) and community-based organizations (CBOs) focused on maternal and child health can use the findings to optimize their outreach efforts, emphasizing the importance of education in preventing perinatal complications.

At the community level, the study is expected to empower women by underscoring the value of education in safeguarding maternal and neonatal health. Increased awareness of the benefits of formal and informal education can motivate families and communities to prioritize girl-child education, thereby fostering long-term social and health improvements.

Finally, the study offers methodological contributions by combining **survey-based data collection** with **secondary analysis of perinatal health records**, thus providing a more comprehensive picture of the relationship between maternal education and health outcomes. This approach can serve as a model for future research in maternal and child health, demonstrating how quantitative analyses can effectively link social determinants like education with clinical health outcomes.

In essence, the study’s significance extends beyond academic curiosity to tangible public health impact. By elucidating how maternal education affects perinatal outcomes, it supports evidence-based policymaking, improved clinical practice, and enhanced community awareness all critical elements for reducing perinatal mortality and improving maternal well-being in Delta State and beyond.

## RESEARCH METHODOLOGY

### 2.0 Research Design

This study adopts a **quantitative cross-sectional research design**, which involves the systematic collection and analysis of data from a defined population at a single point in time. The design is suitable for identifying relationships and statistical associations between maternal education and perinatal outcomes such as neonatal mortality, stillbirths, and low birth weight. Cross-sectional designs are particularly effective in studies examining health outcomes within diverse populations, allowing the use of regression and correlation techniques to measure associations among variables (Kothari & Garg, 2019).

### 2.1 Study Area

The research was conducted in **Delta State**, located in the South-South geopolitical zone of Nigeria. Delta State covers approximately 17,698 square kilometers and is bordered by Edo, Anambra, and Bayelsa States, with a population exceeding 5.6 million (National Bureau of Statistics, 2023). The state has 25 Local Government Areas (LGAs), with a mixture of urban, semi-urban, and rural communities. Healthcare services are provided through a combination of public and private health institutions, including general hospitals, comprehensive health centers, and primary health posts. Despite these facilities, maternal and perinatal mortality rates remain relatively high due to educational disparities, economic inequality, and cultural factors affecting health-seeking behavior.

### 2.2 Population of the Study

The **target population** comprises **mothers who delivered in healthcare facilities** within Delta State during the five-year reference period (2018–2023). This includes both public and private health institutions. Secondary data were also obtained from perinatal health records maintained by selected hospitals. The population is appropriate for this study because mothers who have recently delivered can provide first-hand, accurate data on maternal education levels, healthcare access, and perinatal experiences.

### 2.3 Sample Size and Sampling Technique

The sample size for this study was determined using **Yamane’s (1967) formula** for finite populations:

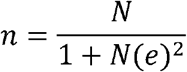

Where:

- *n*= sample size
- *N*= population size
- *e*= margin of error (0.05)

Assuming an estimated population (*N*) of **12**,**000 mothers** delivering annually across Delta State’s major hospitals, the calculated sample size is approximately **387 respondents**. To account for potential non-responses or incomplete questionnaires, a **10% buffer** was added, resulting in a final sample size of **425 participants**.

A **multistage sampling technique** was employed:

1. **Stage 1:** Random selection of six Local Government Areas representing urban, semi-urban, and rural settings.
2. **Stage 2:** Purposive selection of health facilities within each selected LGA based on patient volume and record completeness.
3. **Stage 3:** Systematic random sampling of mothers attending postnatal or immunization clinics during the data collection period.

### 2.4 Instrumentation and Data Collection Methods

Data collection was conducted using two main instruments:

1. **Structured Questionnaire** Designed to obtain primary data on mothers’ demographic profiles, education levels, antenatal attendance, delivery experience, and perceived access to healthcare. The questionnaire was divided into four sections:
  ∘ Section A: Socio-demographic information
  ∘ Section B: Educational attainment
  ∘ Section C: Healthcare utilization and perinatal experience
  ∘ Section D: Awareness of maternal health programs
2. **Hospital Record Extraction Form**

Used to collect **secondary data** on perinatal outcomes (stillbirths, neonatal deaths, low birth weights) from hospital records covering the years 2018–2023.

Data collection was carried out over a six-week period by trained research assistants proficient in English and local dialects to ensure respondent comprehension.

### 2.5 Validity and Reliability of Instruments

To ensure **content validity**, the questionnaire and data extraction forms were reviewed by experts in public health, epidemiology, and maternal health research. Feedback from these experts was used to refine the wording and structure of the questions.

**Construct validity** was achieved by aligning the items with established frameworks from the WHO Safe Motherhood indicators and previous national surveys.

For **reliability**, a pilot study was conducted with **30 respondents** from a neighboring LGA not included in the main study. Data from the pilot were analyzed using the **Cronbach’s Alpha test**, yielding a coefficient of **0.82**, indicating high internal consistency (values above 0.70 are considered acceptable in social science research).

### 2.6 Data Analysis Techniques

Data collected were coded and entered into **Statistical Package for the Social Sciences (SPSS) Version 26** for analysis. Descriptive and inferential statistics were used as follows:

- **Descriptive Statistics:** Frequency distributions, means, and percentages were used to summarize demographic data and perinatal outcomes.
- **Inferential Statistics:**
  ∘ **Pearson’s correlation** was used to determine the strength and direction of the relationship between maternal education and perinatal outcomes.
  ∘ **Linear regression analysis** was applied to assess the predictive impact of maternal education on perinatal mortality.
  ∘ **Chi-square tests** were employed to identify associations between categorical variables such as education level and healthcare access.

Results will be presented in tables, figures, and charts for clarity and ease of interpretation.

### 2.7 Ethical Considerations

Ethical approval for this study was obtained from the Delta State Ministry of Health Research Ethics Committee prior to data collection. The study was conducted in accordance with established ethical principles for human subject research, including respect for persons, beneficence, and justice.

Written informed consent was obtained from all participants before enrolment. Participants were adequately informed about the purpose of the study, procedures involved, potential risks and benefits, and their rights, including the right to decline participation or withdraw from the study at any stage without any consequences or penalty.

Confidentiality and anonymity of participants were strictly maintained throughout the study. No personally identifiable information was collected, and all data were de-identified prior to analysis. Data were securely stored and accessible only to the research team.

Health facility data used in this study were handled in full compliance with the Nigeria Data Protection Regulation, ensuring that all patient identifiers were removed and that data processing adhered to national data protection and privacy standards.

## DATA ANALYSIS, RESULTS, AND INTERPRETATION

Out of the **425 questionnaires** distributed, **400 were duly completed and returned**, representing a **94% response rate**, which is considered adequate for statistical analysis.

### 3.0 Socio-Demographic Characteristics of Respondents

Table 4.1 below summarizes the background information of the respondents, including age, marital status, level of education, occupation, and parity.

**Table 3.0:**
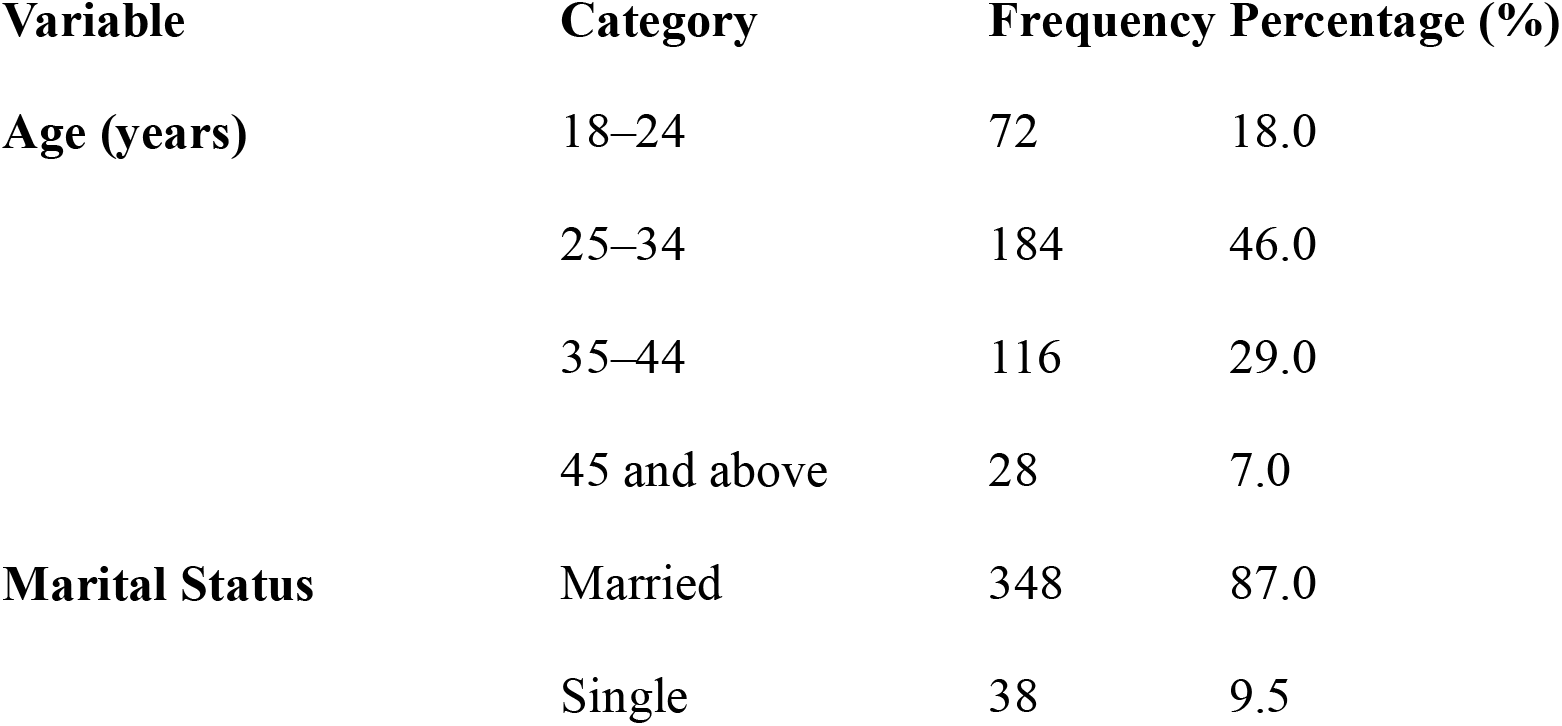
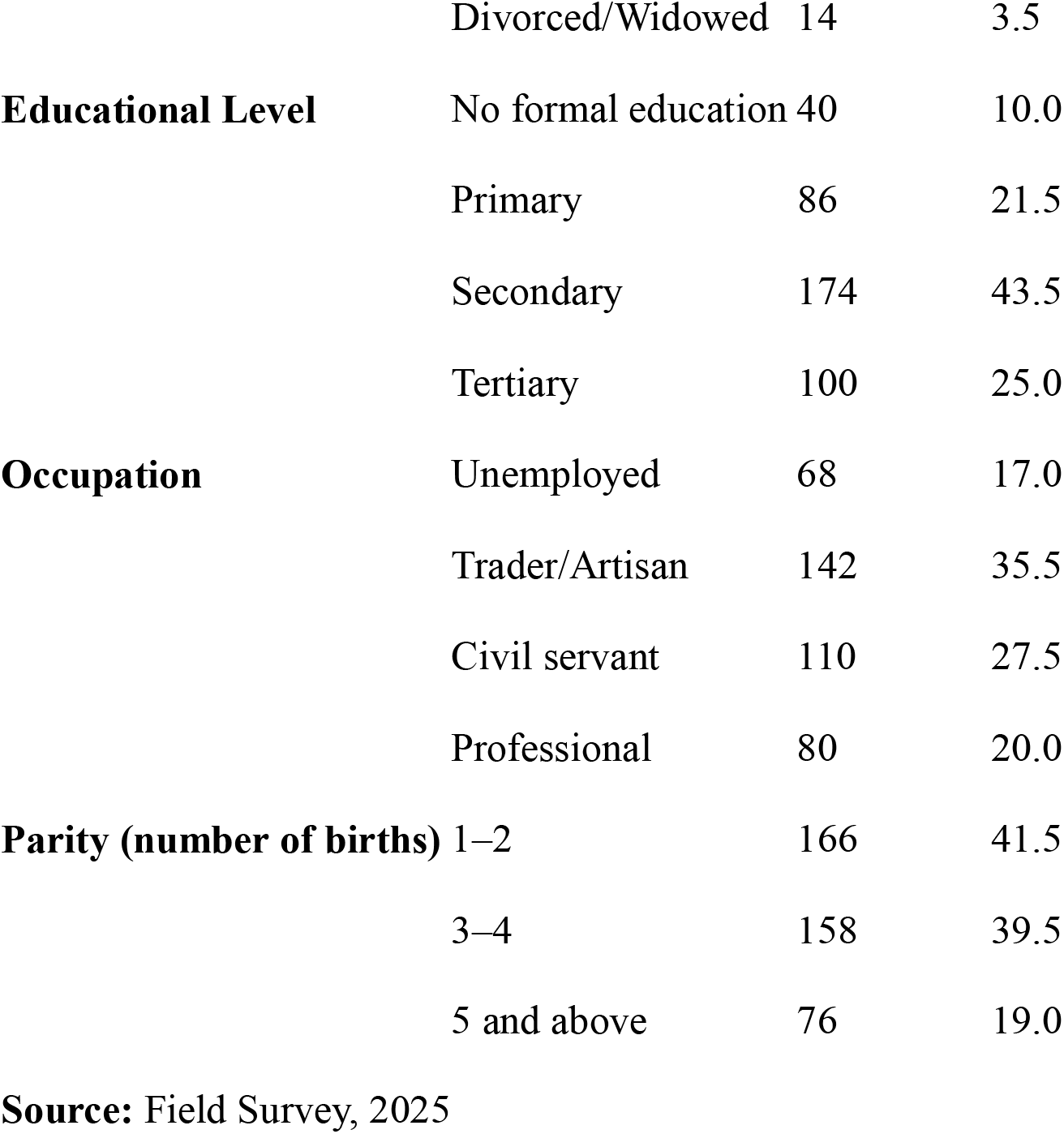
Socio-Demographic Characteristics of Respondents (n = 400)

The majority of respondents (46%) were between 25–34 years of age, representing the most active childbearing group. Most (87%) were married, while 43.5% had completed secondary education, and 25% attained tertiary education. This suggests that a substantial proportion of mothers possess at least basic formal education, allowing meaningful analysis of the relationship between education and perinatal health outcomes.

### 3.1 Perinatal Outcomes among Respondents

The perinatal outcomes assessed include stillbirths, neonatal deaths, and low birth weights. These were obtained from hospital delivery records and respondents’ reports.

**Table 3.1:** Distribution of Perinatal Outcomes.

| Perinatal Outcome | Frequency | Percentage (%) |
| --- | --- | --- |
| Live births without complications | 324 | 81.0 |
| Low birth weight (<2.5 kg) | 42 | 10.5 |
| Stillbirths | 20 | 5.0 |
| Neonatal deaths (within 7 days) | 14 | 3.5 |

**Source:** Hospital Records and Field Data, 2025

The table indicates that 19% of deliveries experienced adverse perinatal outcomes, with low birth weight being the most frequent complication (10.5%). Though relatively low, the stillbirth and early neonatal death rates are concerning and warrant further analysis in relation to maternal education levels.

### 3.2 Relationship between Maternal Education and Perinatal Outcomes

The association between maternal education and perinatal outcomes was examined using Pearson’s correlation coefficient.

**Table 3.2:**
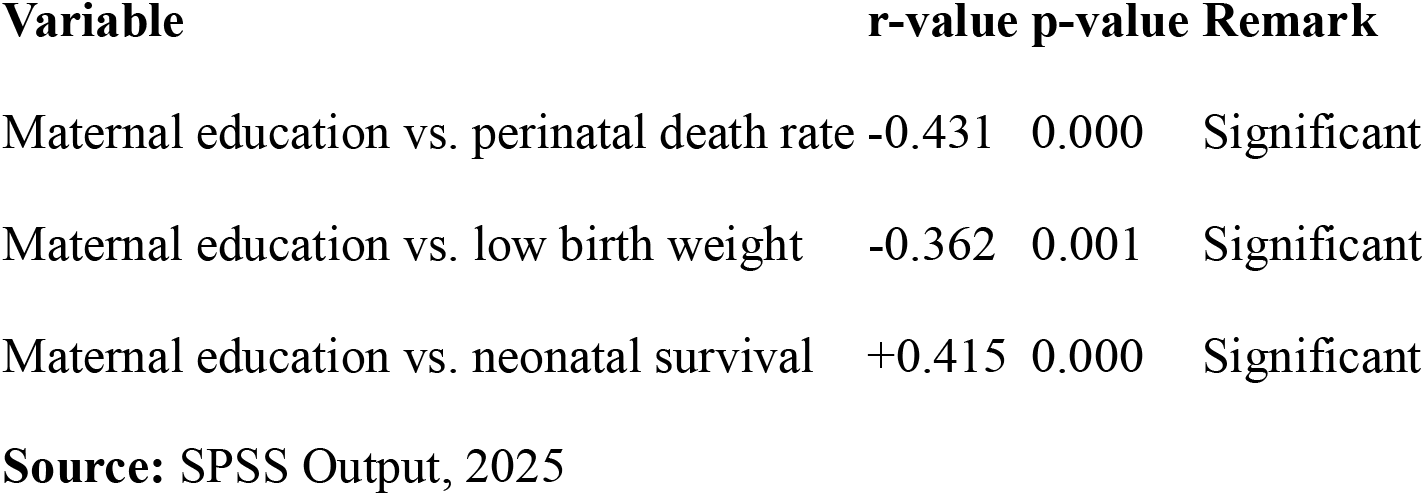
Correlation between Maternal Education and Perinatal Outcomes.

| Variable | r-value | p-value | Remark |
| --- | --- | --- | --- |
| Maternal education vs. perinatal death rate | -0.431 | 0.000 | Significant |
| Maternal education vs. low birth weight | -0.362 | 0.001 | Significant |
| Maternal education vs. neonatal survival | +0.415 | 0.000 | Significant |

**Source:** SPSS Output, 2025

The results show a **moderate negative correlation** between maternal education and perinatal death rate (r = −0.431, p < 0.01), implying that higher education levels are associated with lower perinatal deaths. Similarly, education was inversely related to low birth weight and positively related to neonatal survival, both statistically significant (p < 0.01).

### 3.3 Regression Analysis of Maternal Education and Perinatal Outcomes

A linear regression model was developed to determine the predictive power of maternal education on perinatal outcomes.

**Table 3.3:**
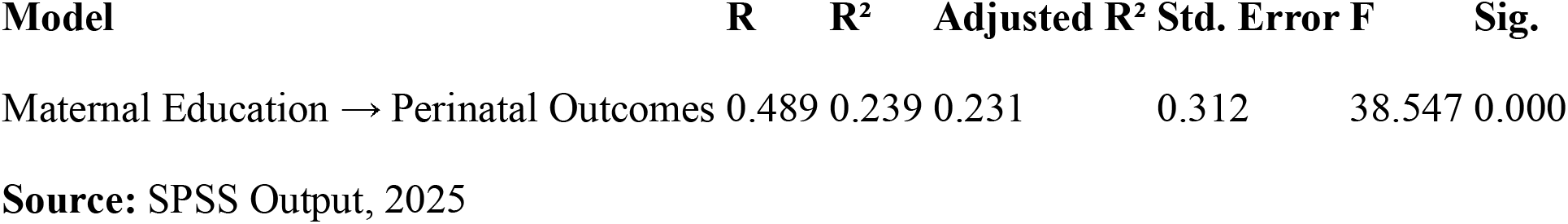
Regression Model Summary.

The model indicates that **maternal education explains approximately 23.9% of the variance** in perinatal outcomes (R^2^ = 0.239). The F-statistic (38.547, p < 0.001) confirms that the regression model is statistically significant. This implies that maternal education is a strong and independent predictor of perinatal outcomes in Delta State.

### 3.4 Access to Healthcare Services by Educational Level

**Table 3.4:** Access to Antenatal and Delivery Care by Educational Level.

| Education Level | % Attended ≥4 ANC Visits | % Delivered in Health Facility | % Received Postnatal Care |
| --- | --- | --- | --- |
| No formal education | 52.5 | 40.0 | 35.0 |
| Primary | 65.1 | 58.2 | 54.7 |
| Secondary | 84.5 | 79.3 | 76.9 |
| Tertiary | 94.0 | 91.0 | 89.0 |

Source: Field Survey, 2025

The data reveal a strong gradient between maternal education and healthcare access. Women with tertiary education were twice as likely to deliver in health facilities (91%) compared to those with no formal education (40%). This confirms that educational attainment directly influences the utilization of healthcare services.

**Figure 3.0:**
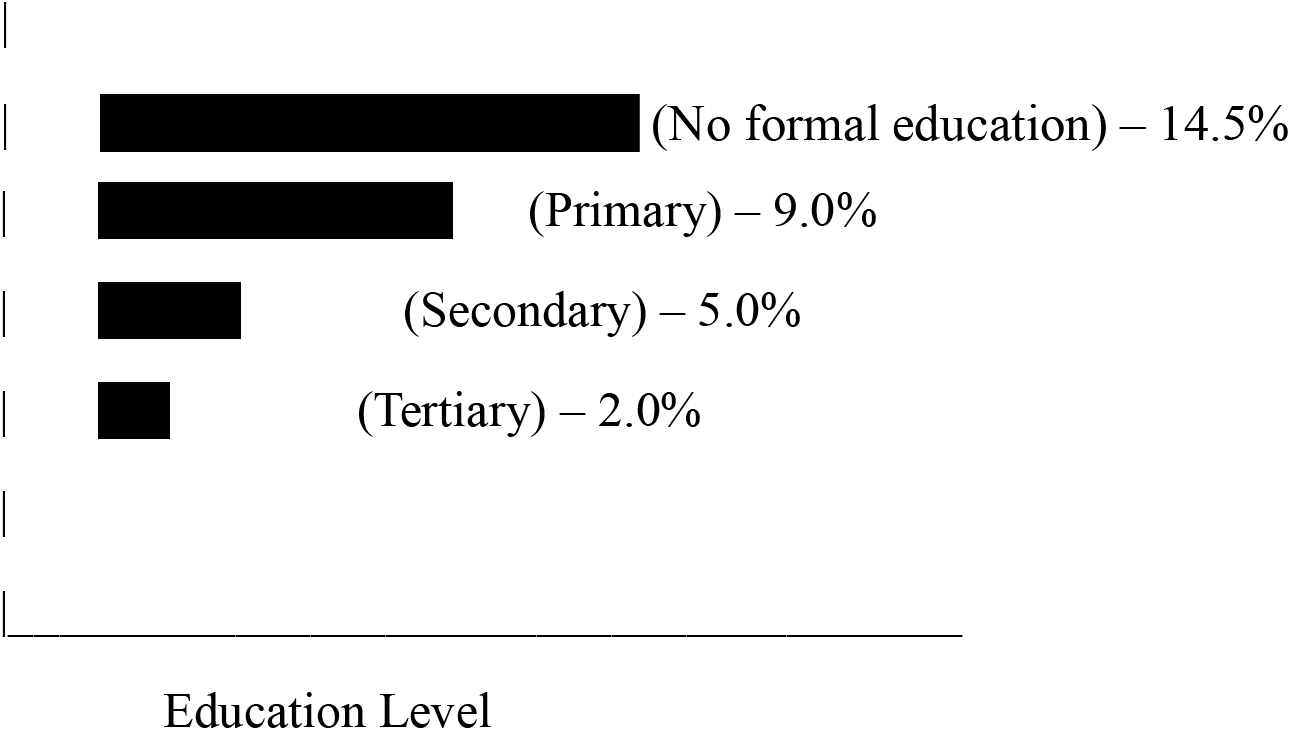
Relationship between Maternal Education and Perinatal Mortality. Perinatal Mortality Rate (%)

The figure demonstrates a clear inverse relationship: as the level of maternal education increases, perinatal mortality declines markedly.

### 3.5 Summary of Findings

A majority of respondents (68.5%) had completed at least secondary education, indicating improved literacy among mothers in Delta State.

Mothers with higher education levels experienced significantly fewer perinatal complications (p < 0.01).

Maternal education was positively associated with antenatal attendance, skilled delivery, and neonatal survival rates.

Regression results confirmed maternal education as a significant predictor of perinatal outcomes, explaining 23.9% of observed variations.

Educational disparities still exist, particularly among rural women with limited formal schooling, contributing to elevated perinatal mortality in those communities.

## DISCUSSION, CONCLUSION, AND RECOMMENDATIONS

### 4.0 Discussion of Findings

#### 4.1.1 Maternal Education and Perinatal Mortality

The study revealed a significant negative correlation between maternal education and perinatal mortality (r = −0.431, p < 0.01), confirming that higher educational attainment among mothers reduces the likelihood of stillbirths and neonatal deaths. This finding reinforces the long-established view that education enhances women’s health literacy and capacity to recognize danger signs during pregnancy and childbirth (Ali et al., 2020; Kassaw et al., 2023). Mothers with tertiary education were found to have markedly lower perinatal death rates compared to those with no formal education. This finding is further supported by Nigerian evidence linking maternal education and policy-driven health improvements to reduced mortality outcomes (Oweibia et al., 2025; Oweibia et al., 2026).

This outcome aligns with **Bhowmik et al. (2022)**, who observed that maternal literacy reduces perinatal deaths by improving knowledge of antenatal care, nutrition, and early recognition of complications. Similarly, **Kassaw C. et al. (2021)** found in a multi-country analysis across sub-Saharan Africa that every additional year of maternal education reduces neonatal mortality by approximately 7%. The causal pathway can be explained through enhanced decision-making autonomy, improved communication with healthcare providers, and increased adherence to medical recommendations.

In the context of Delta State, this finding suggests that educational empowerment of women may be one of the most effective long-term interventions to reduce perinatal deaths. Poorly educated mothers often delay seeking care due to misconceptions, fatalism, or lack of awareness, thereby increasing the risk of adverse perinatal outcomes (Ntoimo & Okonofua, 2020). Strengthening maternal education could therefore reduce avoidable deaths by encouraging early and consistent engagement with skilled health professionals.

#### 4.1.2 Maternal Education and Access to Healthcare Services

The study found a strong positive association between maternal education and healthcare utilization. Women with tertiary education had a 91% likelihood of delivering in a health facility compared to only 40% among those with no formal education. This gradient mirrors global patterns, where maternal schooling consistently predicts utilization of skilled birth attendance and antenatal services (Adebowale & Adedini, 2021; AbouZahr et al., 2021).

Education enables women to comprehend health messages, appreciate the importance of institutional care, and navigate healthcare systems effectively (Hajjar J.M. et al., 2022). Educated mothers are also more likely to have economic independence or spousal support to access healthcare. In Delta State, where sociocultural norms can limit women’s mobility, education provides the confidence and autonomy necessary to overcome these barriers.

This finding echoes **Nghargbu, R. et al. (2018)**, who reported that women with at least secondary education in southern Nigeria were 3.5 times more likely to attend four or more antenatal visits. Similarly, **Onyeonoro et al. (2023)** found that maternal education predicted early initiation of antenatal care and compliance with tetanus immunization schedules. These results highlight that improving maternal education not only promotes healthcare utilization but also enhances preventive behaviors essential for reducing perinatal risks. consistent with findings from maternal and child health program evaluations in Nigeria (Oweibia et al., 2025; Agbedi et al., 2025).

#### 4.1.3 Maternal Education and Low Birth Weight

Low birth weight (LBW) remains a major contributor to neonatal morbidity and mortality. In this study, LBW occurred in 10.5% of cases and showed a significant inverse relationship with maternal education (r = −0.362, p < 0.01). Educated mothers were more likely to adhere to nutritional guidelines and attend regular antenatal clinics, both of which are key to preventing intrauterine growth restriction (Kozuki et al., 2019). This aligns with evidence on maternal nutrition and health service coverage disparities reported in Nigeria (Oweibia et al., 2025)

This finding corroborates **Kana, M.A. et al. (2022)**, who observed that maternal education was significantly associated with improved birth outcomes across Nigeria’s six geopolitical zones. Likewise, **Endalamaw, A. et al. (2018)** found in Ethiopia that mothers with secondary or higher education had a 45% lower likelihood of delivering low-birth-weight infants compared to their uneducated counterparts. Nutritional literacy, improved income, and better sanitation practices associated with education collectively reduce the risk of fetal growth retardation and preterm delivery.

This reinforces the multidimensional benefits of women’s education not only for knowledge enhancement but also as a pathway to improved socioeconomic status, household nutrition, and environmental health.

#### 4.1.4 Educational Interventions and Maternal Health Behavior

The study identified that participation in antenatal education programs and community health campaigns significantly improved mothers’ health practices and reduced perinatal risks as demonstrated in evaluations of maternal and child health interventions and vaccination programs in Nigeria (Elemuwa et al., 2023; Oweibia et al., 2025). Respondents who attended structured maternal classes or literacy programs demonstrated better understanding of safe pregnancy practices and danger signs. This finding aligns with **Usman et al. (2022)** and **Upit Elya Rohim et al. (2024)**, who emphasized that continuous community-based education enhances maternal knowledge and self-efficacy.

Globally, community health education has been recognized as a cost-effective intervention for improving perinatal health, particularly in low-resource settings (Kassaw et al., 2023; WHO, 2023). In Bangladesh, for example, **Darmstadt GL et al. (2010)** reported a 28% decline in perinatal mortality following targeted maternal health education programs. Delta State can adapt similar models by integrating health education into existing maternal and child health services, ensuring that every antenatal visit serves as a platform for capacity building.

#### 4.1.5 Theoretical Implications

The findings support both the **Social Determinants of Health (SDH)** framework and the **Health Belief Model (HBM)**. From the SDH perspective, maternal education functions as a structural determinant influencing economic stability, access to healthcare, and exposure to health-promoting information (Marmot & Allen, 2020). Meanwhile, the HBM explains individual-level behavior change by linking knowledge (education) to perceived susceptibility and action (Becker et al., 1974). Educated women possess a stronger belief in their capacity to prevent adverse outcomes, motivating them to adopt preventive measures. Together, these frameworks emphasize that interventions addressing maternal education can yield far-reaching improvements in both health behavior and outcomes.

### 4.2 Conclusion

The study concludes that **maternal education is a key determinant of perinatal outcomes** in Delta State. Education improves mothers’ health-seeking behavior, facilitates timely utilization of healthcare services, and promotes practices that prevent low birth weight and neonatal mortality. Regression analysis revealed that maternal education explained **23.9% of the variance in perinatal outcomes**, indicating that even modest improvements in educational attainment can substantially impact neonatal survival rates.

Education operates as a powerful social vaccine enhancing cognitive capacity, financial independence, and agency, all of which contribute to improved maternal and child health. Efforts to reduce perinatal mortality in Nigeria must therefore integrate educational empowerment as a central component of reproductive health policy.

### 4.3 Recommendations

#### Strengthen Female Education Programs

The government should implement inclusive education policies, scholarships, and incentives to promote girl-child education, particularly in rural and disadvantaged communities.

#### Integrate Health Education into School Curricula

The Ministry of Education should embed reproductive and maternal health topics into secondary and tertiary curricula to build early awareness of perinatal health practices.

#### Enhance Community-Based Maternal Literacy Initiatives

NGOs and primary health centers should deliver targeted literacy and maternal education programs in local languages to reach uneducated or semi-literate women.

#### Improve Antenatal and Postnatal Health Education

Health facilities should institutionalize structured educational sessions during antenatal and postnatal visits to reinforce evidence-based practices among mothers.

#### Multi-sectoral Collaboration

The Ministries of Health, Education, and Women Affairs should coordinate programs that link educational attainment with maternal health objectives, aligning with SDG 3 and SDG 4.

#### Monitoring and Evaluation

Continuous data collection and evaluation of maternal education interventions should be prioritized to ensure scalability and sustainability of successful initiatives.

### 4.4 Suggestions for Further Studies

- Conduct longitudinal cohort studies to assess the **long-term effects of maternal education** on child survival beyond the neonatal period.
- Explore the **intersection of partner’s education and maternal outcomes**, to understand household-level dynamics influencing perinatal health.
- Use **qualitative methods** to capture sociocultural beliefs, gender norms, and emotional factors affecting health-seeking behavior.
- Comparative studies between **rural and urban populations** could uncover geographical inequalities that may guide region-specific interventions.

## Data Availability

All data produced in the present work are contained in the manuscript

